# Barriers and facilitators to women general practitioners’ career progression: a systematic review

**DOI:** 10.1101/2023.09.26.23296133

**Authors:** Laura Jefferson, Elin Webster, Su Golder, Katie Barnett, Nicola Greenwood, Veronica Dale, Karen Bloor

## Abstract

**Objective:** To identify barriers and facilitators to women GPs’ career progression.

**Design:** Systematic review of qualitative and quantitative studies.

**Setting:** Studies conducted in the UK NHS general practice setting.

**Participants:** General practitioners.

**Main outcome measures:** Barriers and facilitators mapped thematically to the COM-B model, including Capability, Opportunity and Motivation as influencers of Behaviour.

**Results:** 21 articles were included in this review, with varied study designs. No relevant intervention studies were identified. There was a lack of recent research evidence; over half were conducted over 20 years ago. Most studies met quality criteria, though there were some problems with reporting and adjustment for potential confounders. Barriers at personal, socio-cultural and system levels were found that inhibit women GPs’ career progression. While some positive changes have been documented across studies that span some thirty years, many challenges remain.

**Conclusion:** Despite general practice being one of the more female-dominated medical specialties, barriers at personal, socio-cultural and system levels continue to inhibit women GPs’ careers. The COM-B model of behaviour change was used to group thematic findings according to the barriers women may face in terms of their capabilities, opportunities and motivations and identify potential policies that could be evaluated as options to support women GPs’ career progression.

## Introduction

Women constitute 52% of full-time equivalent general practitioners (GPs) in England (1). A recent independent review of the gender pay gap in medicine identified general practice as the specialty with the greatest gender pay gap (33.5% unadjusted), one of the highest of any UK profession(2). While differences in working hours, age and experience account for approximately half of this variation, the lower rate at which women GPs become partners (or ‘principals’) is also likely to be a significant factor since it is associated with higher pay and profit-sharing (2). Women currently comprise 41% of UK GP partners (1) and the presence of a ‘glass ceiling’ in medicine has been widely described, referring to women’s lower ability to progress in their careers and worse reported pay and conditions (3-7).

Studies exploring gender differences in medical careers tend to focus on hospital specialities, particularly those with historically lower proportions of women, such as surgery, where differential treatment and ‘old boys’ clubs’ have been shown to discriminate against women doctors (8-10). Hafferty (11) described a ‘hidden curriculum’ of cultural norms and customs in medical institutions some 25 years ago, but a recent BMA report on sexism in medicine highlights a worryingly persistent negative culture in today’s medical system: 91% of women doctors reported experiencing sexism at work (12).

The impact of wider societal gendered expectations creates differential pressure on women doctors’ caring responsibilities, even in dual doctor marriages (13). Evidence from international primary care settings recently suggested this societal expectation places additional pressure on women GPs’ life transitions (14). In the UK, recent research is lacking on this topic and the wide gender pay gap in general practice (2) highlights a need to explore the barriers and facilitators that influence women GPs’ career progression. As part of a wider UK policy research project, we undertook a systematic review of the existing UK evidence to identify evidence gaps and synthesise evidence, highlighting potential avenues for intervention development that may support women GPs’ careers.

## Method

We used systematic review methods, following the Cochrane guidelines for conducting systematic reviews (15) and, to ensure transparency of reporting, we used a PRISMA checklist (16). To reduce potential duplication of effort, we registered the study in advance (PROSPERO ID: CRD42023384176).

### Search strategy

We employed a varied search strategy, using both database searching and wider sources to search for reports. Our sources included MEDLINE, Embase and the Healthcare Management Information Consortium (HMIC) database (initial search 5^th^ January 2022, repeated 4^th^ January 2023), alongside searches of Google Scholar, key websites, reference lists and online e-theses (via EThOs) to capture grey literature. See Supplementary File A for full search strategy. Forward and backward citation searching was conducted on included studies. No date or language restrictions were applied.

### Inclusion criteria

Studies were included if they investigated barriers and facilitators of career progression, including uptake of partnership roles, for women general practitioners. Included studies were either those exploring specifically the experiences of women, or drawing comparisons across genders. We excluded studies of multiple health professional groups if GP findings were not disaggregated. Since this study was embedded within a wider UK policy research project, we focused on studies conducted in the UK, excluding non-UK studies. No limits were applied according to study design, but we included only empirical research evidence, excluding case reports and editorials.

### Selection of studies

We downloaded search results into Covidence (17) to de-duplicate and conduct screening. Two of five reviewers independently completed initial screening of titles and abstracts, followed by full text screening. We resolved any disagreements between reviewers through discussion or a third reviewer (LJ or SG).

### Data Extraction and Quality Assessment

We used a pre-piloted data extraction form, with one of four reviewers extracting data and cross checking a 20% sample to ensure consistency. Depending on the study design, we used the Joanna Briggs Institute (JBI) Checklist for Analytical Cross-Sectional Studies (18) or the Critical Appraisal Skills Programme (CASP) checklist tool for qualitative studies (19) for quality assessment. Two reviewers independently performed quality appraisal, with arbitration by a third reviewer in cases of disagreement (5%). Studies were not excluded based on quality.

### Data Synthesis

To summarise the study findings we used narrative synthesis, as variation across studies prohibited the use of quantitative approaches. We managed and sorted data in MS Excel and then employed thematic qualitative synthesis to map findings using the COM-B theoretical model of behaviour change (20). This provided a structured approach to identify barriers to behaviour, acknowledging both individual and contextual factors that may affect an individuals’ likelihood of engaging in behaviours that promote career progression, for example applying for a partnership role (20). The *capability (C)* construct refers to an individuals’ psychological and physical (personal) capabilities, while the *opportunity (O)* construct relates to environmental, social and physical opportunities (20). Together, factors relating to capability and opportunity factors are expected to influence the relationship between *motivation (M)* and *behaviour (B)*, whereby motivation relates to an individuals’ beliefs, values, feelings, confidence and intentions towards a behaviour (20).

We used an iterative process, moving through the stages of initial ‘free coding’ to more descriptive and then later, analytical themes using the overarching themes of ‘Capability’, ‘Opportunity’ and ‘Motivation’(21). Each stage was undertaken with regular consultation and discussion between researchers who had methodological and topic expertise, some of whom also had lived experience as female doctors.

## Results

### Search results

In total, we identified 2356 studies from databases and grey literature searching. After removal of duplicates, 1306 articles were screened as titles and abstracts. We excluded 1017 at this initial stage, leaving 289 for full text review. 21 studies met the inclusion criteria and were included in this review (Figure 1) (12, 22-41).

**Figure 1:**
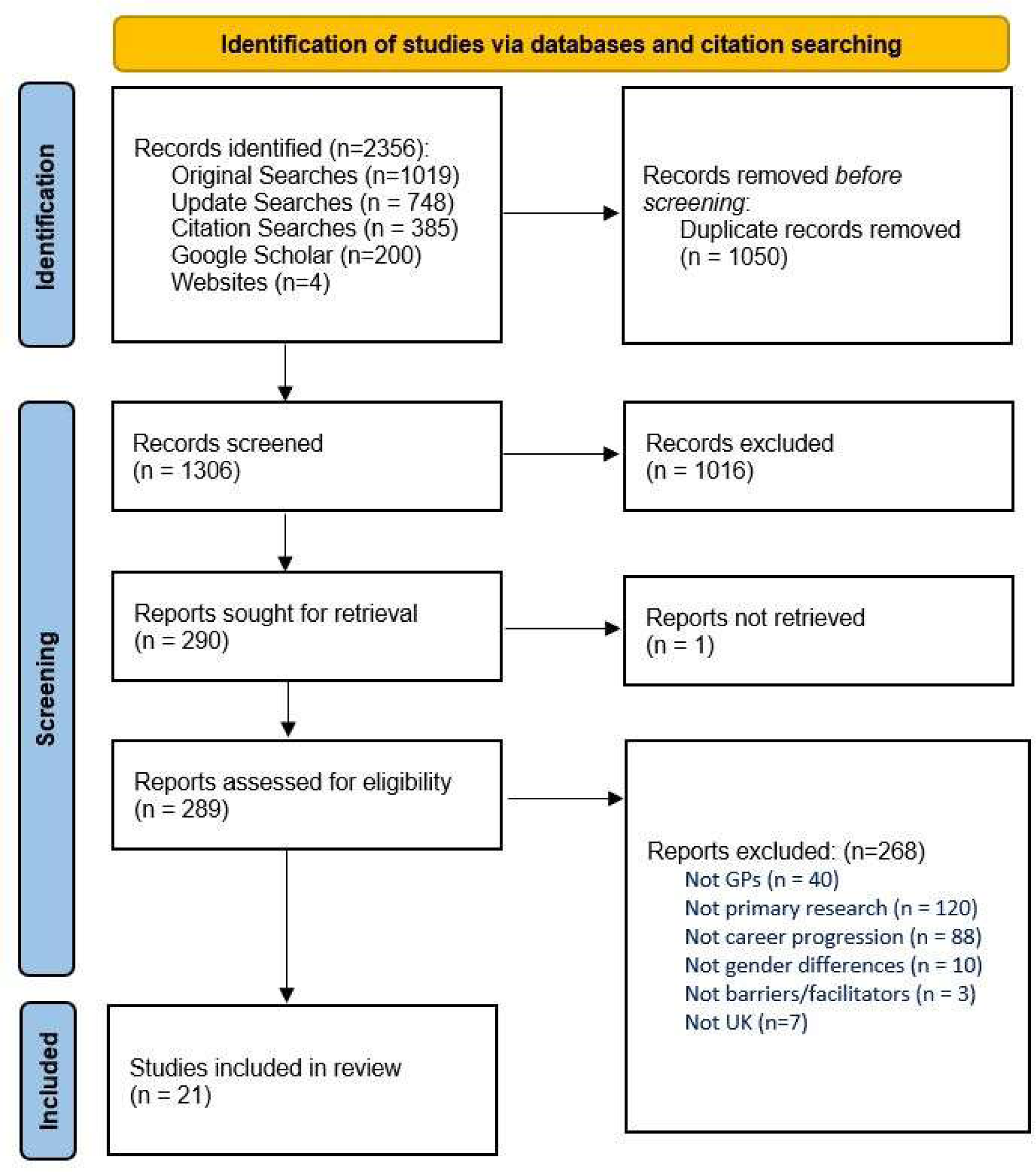
PRISMA diagram.

### Study Characteristics

Study designs varied, with 10 cross-sectional surveys, six qualitative interview studies, two secondary econometric analyses, two mixed methods studies and one discrete choice experiment. We found no relevant intervention studies.

The majority of studies were conducted some time ago; more than half were over 20 years ago and only three studies were conducted in the last ten years (12, 25, 31). Of these, one was a PhD thesis that only included four GPs (31). Studies were geographically dispersed across the UK, with five UK-wide, three in England, three in Scotland, one England & Wales and seven in single-locations within the UK.

Six studies include only women, while the remaining 15 studies explored gender differences. Sample sizes ranged from a qualitative study with four GPs to an econometric analysis of 2,271 GPs (median 368).

### Quality Assessment

The quality of studies was generally good, with all providing valuable insights (Table 2 and 3). Though all but one cross-sectional study identified potential confounding factors, only 6/13 used strategies for dealing with such confounders, for example through statistical analyses. All other components of the quality assessment of cross-sectional studies were generally good. Qualitative studies were generally sound, though one study conducted in 1989 was rated as ‘unclear’ or inadequate across numerous categories (38). Qualitative studies tended not to reflexively consider relationships between researchers and participants and only two described ethical considerations (25, 31). Insufficient detail about analysis hindered quality assessment in three studies (29, 38, 41).

**Table 1:**
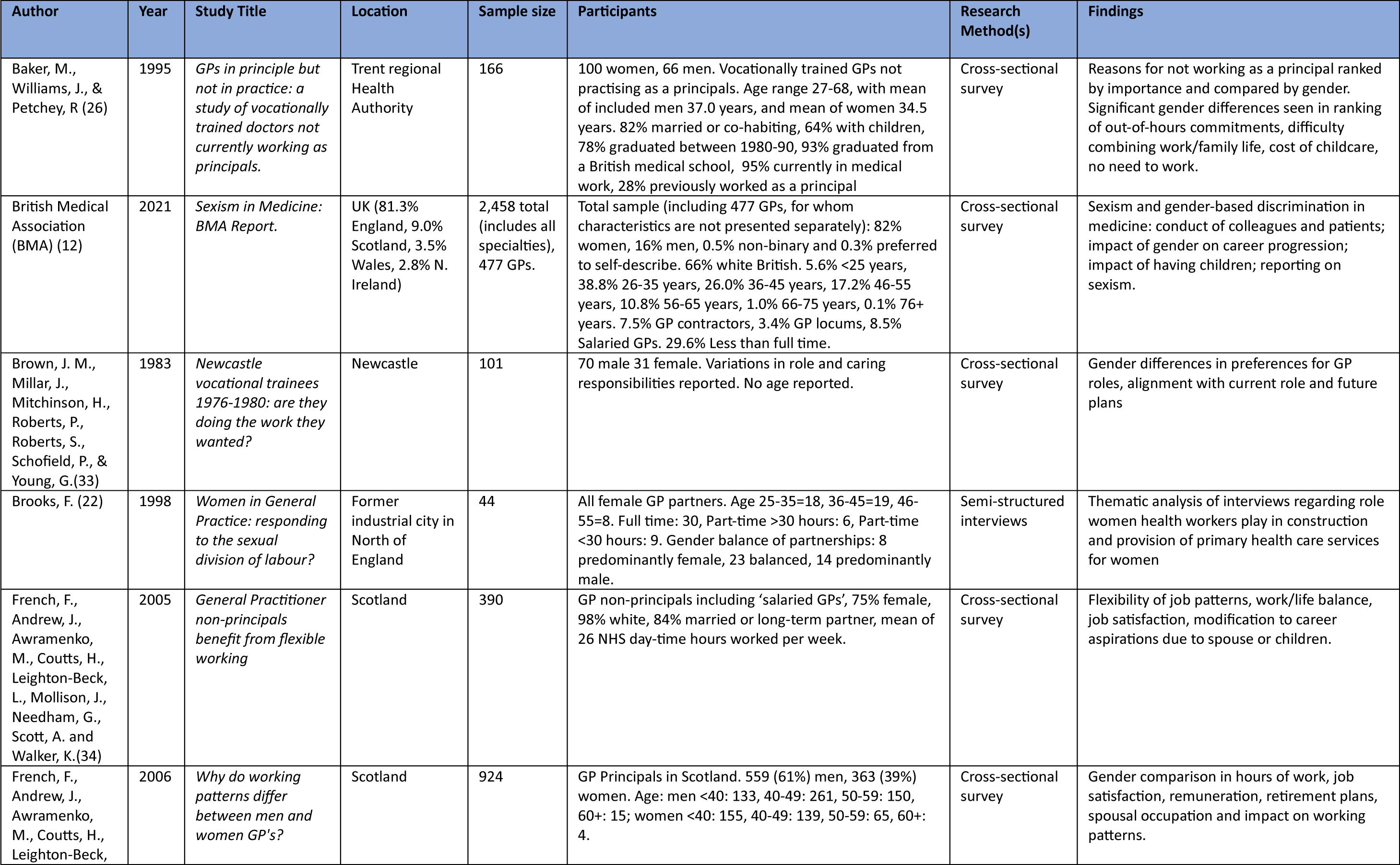
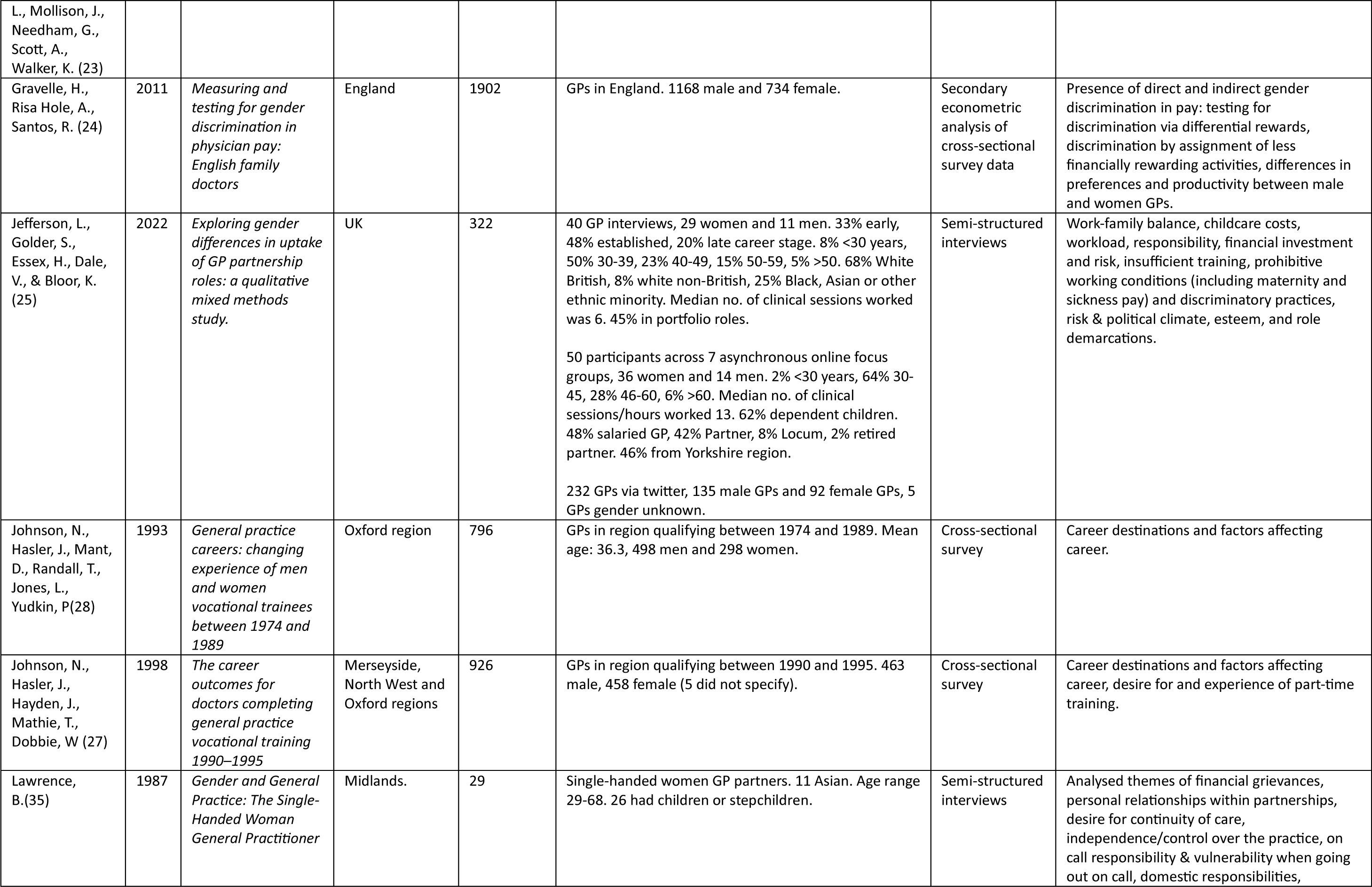
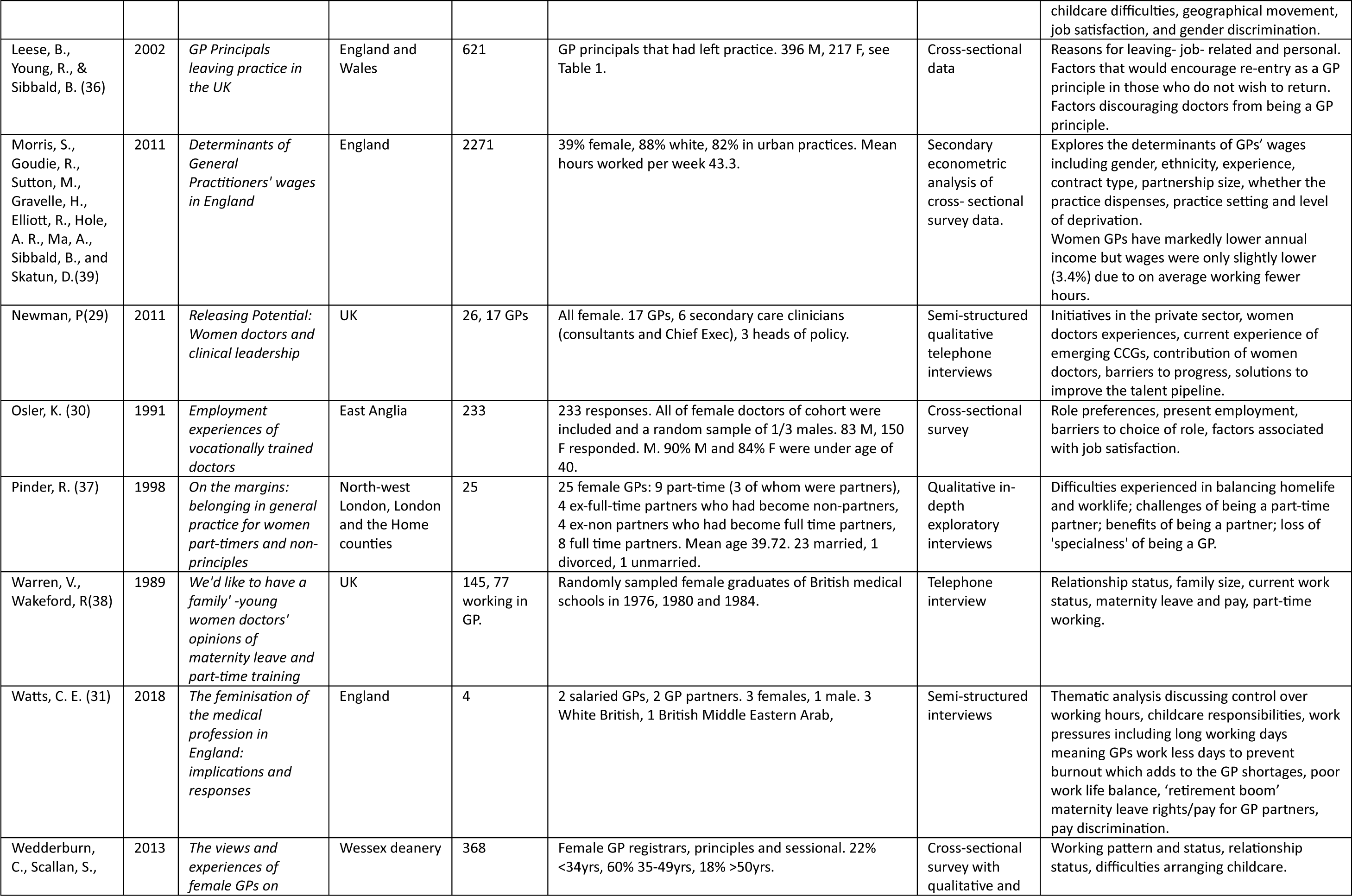
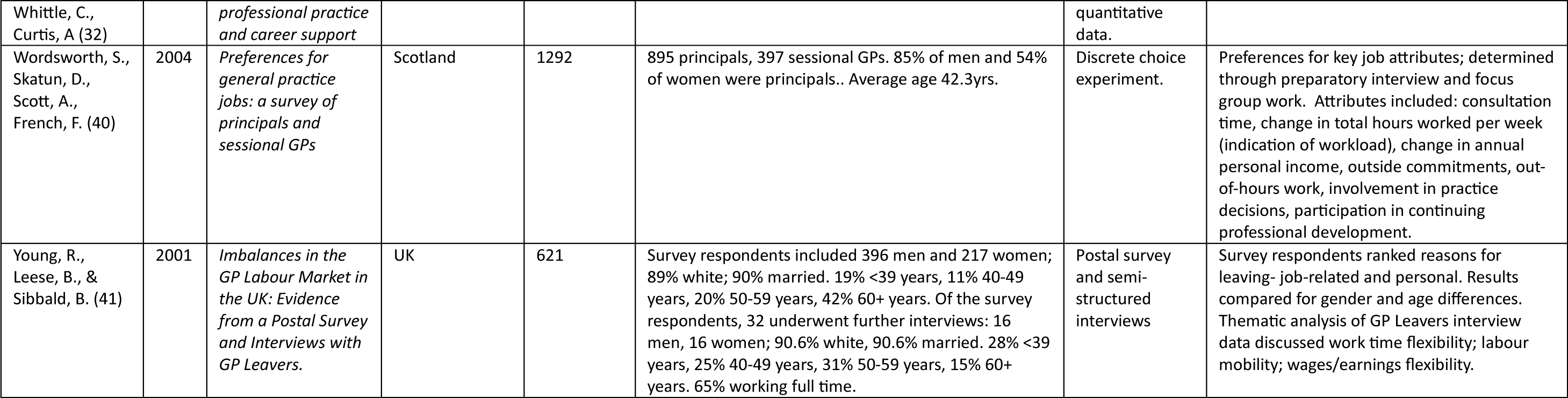
Characteristics of included studies.

**Table 2:** Quality Appraisal of cross-sectional studies.

| Author (Year) | 1. Were the criteria for inclusion in the sample clearly defined? | 2. Were the study subjects and the setting described in detail? | 3. Was the exposure measured in a valid and reliable way? | 4. Were objective, standard criteria used for measurement of the condition? | 5. Were confounding factors identified? | 6. Were strategies to deal with confounding factors stated? | 7. Were the outcomes measured in a valid and reliable way? | 8. Was appropriate statistical analysis used? |
| --- | --- | --- | --- | --- | --- | --- | --- | --- |
| Brown et al., 1983 | Yes | Unclear | Yes | Yes | Yes | No | Yes | No |
| French et al., 2006 | Yes | Yes | Yes | Yes | Yes | Yes | Yes | Yes |
| Gravelle et al, 2011 | Yes | Yes | Yes | Yes | Yes | Yes | Yes | Yes |
| Johnson et al., 1993 | Yes | Yes | Yes | Yes | Yes | No | Yes | Yes |
| Johnson et al., 1998 | Yes | Yes | Yes | Yes | No | No | Yes | Yes |
| Leese et al, 2002 | Yes | Yes | Yes | Yes | Yes | Yes | Yes | Yes |
| Osler, 1991 | Yes | Yes | Yes | Yes | Yes | No | Yes | Yes |
| Wedderburn et al., 2013 | Yes | Yes | Yes | Yes | Yes | No | Yes | Yes |
| Wordsworth et al, 2004 | Yes | Yes | Yes | Yes | Yes | Yes | Yes | Yes |
| Baker et al, 1995 | Yes | Yes | Yes | Yes | Yes | No | Yes | Yes |
| French et al., 2005 | Yes | Yes | Yes | Yes | Yes | No | Yes | Yes |
| Morris et al., 2010 | Yes | Yes | Yes | Yes | Yes | Yes | Yes | Yes |
| BMA, 2021 | Yes | Unclear | Yes | Yes | Yes | Yes | Unclear | Yes |

### Thematic findings

The COM-B model of behaviour (20) was used to group thematic findings according to the barriers women may face in terms of their capabilities, opportunities and motivations – all influencing their likelihood to adopt behaviours relating to career progression. This model, and the corresponding sub-themes are outlined in Figure 2, summarised below (see Table 4 for further detailed findings).

**Figure 2:**
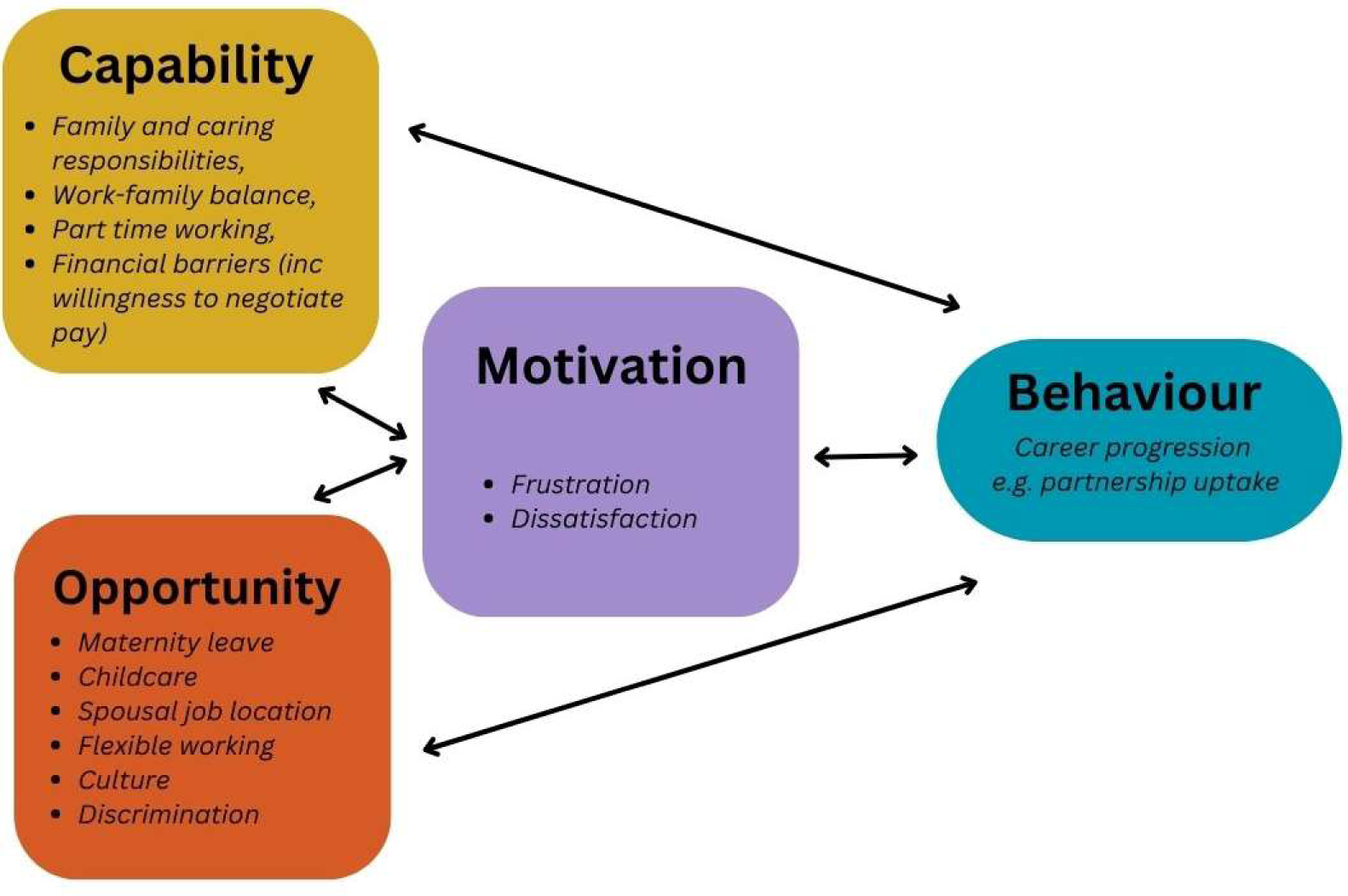
COM-B model and gendered barriers to career progression.

**Table 3:** Quality Appraisal of qualitative studies.

| Author (year) | Section A: Are the results valid? |  |  |  |  |  | Section B: What are the results? |  |  | Section C: Will the results help locally? |
| --- | --- | --- | --- | --- | --- | --- | --- | --- | --- | --- |
|  | 1. Was there a clear statement of the aims of the research? | 2. Is a qualitative methodology appropriate? | 3. Was the research design appropriate to address the aims of the research? | 4. Was the recruitment strategy appropriate to the aims of the research? | 5. Was the data collected in a way that addressed the research issue? | 6. Has the relationship between researcher and participants been adequately considered? | 7. Have ethical issues been taken into consideration? | 8. Was the data analysis sufficiently rigorous? | 9. Is there a clear statement of findings? | 10. How valuable is the research? |
| Brooks, 1998 | Yes | Yes | Yes | Yes | Yes | Unclear | Unclear | Yes | Yes | Valuable |
| Newman, 2011 | Yes | Yes | Yes | Unclear | Yes | Unclear | Unclear | Unclear | Yes | Valuable |
| Pinder, 1998 | Yes | Yes | Yes | Yes | Yes | Unclear | Unclear | Yes | Yes | Valuable |
| Warren & Wakeford, 1989 | No | Yes | No | Unclear | Unclear | Unclear | Unclear | Unclear | Yes | Valuable |
| Jefferson et al., 2022 | Yes | Yes | Yes | Yes | Yes | Yes | Yes | Yes | Yes | Valuable |
| Young et al., 2001. | Yes | Yes | Yes | Yes | Yes | Unclear | Unclear | Unclear | Yes | Valuable |
| Watts, 2018 | Yes | Yes | Yes | Yes | Yes | Yes | Yes | Yes | Yes | Valuable |
| Lawrence, 1987 | Yes | Yes | Yes | Yes | Yes | Yes | Yes | Yes | Yes | Valuable |

**Table 4:** Further detailed findings.

|  |  |
| --- | --- |
| Capability | **Reasons for differential earnings** included women being less likely to negotiate pay (25), women experiencing financial exploitation in group practices with male senior partners (35), women being less motivated by financial gain (25, 35) and lack of financial security in partnerships when taking maternity leave (25, 31, 38). At a personal level, these challenges related to the personal cost of taking time off for maternity leave, but also paying locum cover for GP partners (25, 31, 38).<br>Jefferson (25) described women's lower comparative earnings due to maternity leave and part-time work resulting in reduced financial incentive to progress careers in comparison to spouses, particularly those in dual-doctor marriages. Knowledge of the New to Partnership Payment Scheme to encourage uptake of partnership roles through a financial incentive of £20,000 to new partners has been described as insufficient amidst wider uncertainty within the profession (25). |
| Opportunity | Several studies described specific challenges around taking **maternity leave**. Women GPs exited training pathways due to 'unwritten rules' around maternity leave during training (30, 38), historical contractual arrangements restricted entitlement to maternity leave (38) and women reported a lack of support or outright negativity from colleagues when broaching maternity leave (12, 37). Financial insecurity associated with maternity leave was highlighted across studies despite large time lags and, presumably, cultural shifts in the workplace (25, 31, 38).<br>**Childcare costs** were a barrier in two studies. While Young (41) found salaried contracts were preferable due to fixed incomes simplifying the financial planning for childcare costs (41). Leese et al (36) suggest that improved childcare and term-time contracts may encourage re-entry to partnerships.<br>Six studies cited **spousal job location** as a barrier to progression. Gender differences were seen in the influence of spouse's work location determining job role (33); in likelihood of leaving a partnership role (36); and in reducing career progression and aspirations (23, 27, 34).<br>In terms of **barriers due to working hours**, out of hours working presented challenges due to safety concerns during overnight home-visits and challenges obtaining out-of-hours childcare at short notice, or reliance on spouse or wider family during these times (35). Wordsworth's discrete choice experiment (40) which asked salaried and partnered GPs about their preferences for hypothetical job components, found **flexibility of hours** associated with salaried roles to be a priority in career choices. Similar findings were reported in a pilot discrete choice experiment by Jefferson (25), with both men and women GPs indicating a preference for flexible working to promote partnership uptake. In studies comparing men and women, inflexible work hours were perceived as a greater barrier to progression by women, with significant gender differences recorded in three studies (27, 28, 36). |
| Motivation | In terms of **satisfaction in career roles**, Osler reported more women than men (39% vs 24%) were working in a role that was not their original choice of work (30). Jefferson (25) found women GPs held more responsibility for supporting teams – further increasing workload. Johnson (28) reported other potential reasons for dissatisfaction, including longer time to become a GP principal for women compared to men and frustration related to the socio-cultural barriers women face in the workplace. |

### Capability: family and finances

Studies historically focused on individuals’ personal circumstances inhibiting capability for career progression – primarily the challenges associated with balancing family and work lives, but also financial barriers. Sixteen of the included studies outlined issues relating to family responsibilities for women GPs, citing greater family commitments as a reason for not pursuing principal roles, difficulties establishing work-family balance and challenges of working full-time. Attitudes were perceived as shifting (25, 31, 35, 41), though recent research shows gendered barriers are clearly still associated with caring responsibilities (25). Longitudinal cohorts reported lowering impact of childcare responsibilities on women doctors’ careers over the life course (27, 28), but almost half of women over 50 still reported childcare challenges (32) and caring responsibilities for adult dependents (41).

Financial barriers to career progression were raised by women GPs in seven studies, often focusing on their lower comparative earnings to men but also culturally gendered barriers including willingness to negotiate pay. Statistically and economically significant lower incomes for women GPs were reported and unexplained by observable characteristics (24, 34, 39). Possible reasons are described in Table 4.

### Opportunity: system issues, culture and discrimination

Socio-cultural and systemic barriers to career progression were found across studies, relating to maternity leave practices including ‘unwritten rules’ and contractual challenges, spousal job location (23, 25, 27, 33, 34, 36), childcare costs (25, 26), flexibility of roles (25-28, 35, 36, 40), cultural challenges within general practice and also overt discrimination (12, 22, 25, 28, 29, 35, 37, 38) (Table 4).

Ten studies discussed flexibility in working hours as a barrier to career progression for women GPs. Prior to 2004, GP partners were personally responsible for providing or organising a 24/7 service for patients (42). Several studies conducted pre-2004 cited out-of-hours working as a barrier to working as a GP partner (26, 36, 40). Flexibility in hours was a priority in Discrete Choice Experiments about career preferences (25, 40) and women GPs were statistically significantly more likely to report inflexible hours as a career barrier than men GPs (27, 28, 36). Recently, part-time or salaried roles were described as increasingly being used to cope with challenging working lives and reduce burnout (25, 31). Flexible working encouraged re-entry to principal posts (36) or after temporary exit e.g. through ‘ramp on and off schemes’ (29) and may encourage later retirement (32).

While strong role models promoted positive workplace cultures (25, 29), discriminatory cultures included negative views of part time working (29, 37); increasing demarcations between salaried and partner GPs (25); stereotypical gendered roles (25); and societal expectations of a doctor being male (22, 35). Studies gave accounts of this being displayed through women’s voices not feeling heard (25, 29), passive lack of support (29), differential treatment and respect from support staff (12, 25, 35), reduced opportunities for leadership roles (12, 29), discriminatory interview practices (12, 38) and historical marginalisation and exclusionary behaviours (22, 28, 29). Only 20% of GPs reported never experiencing sexism, though faring better than doctors overall (of whom only 1% had never experienced sexism), this bar is set very low (12).

### Motivation: frustration in roles

Barriers related to an individual’s motivations to progress their careers were less explicitly discussed in studies, but rather inferred due to the likelihood of personal capabilities and wider socio-cultural opportunities impacting individual’s confidence and beliefs around career progression. Women GPs described frustration with being given a higher burden of ‘women’s work’ – particularly caseloads relating to women’s, children’s, and mental health as a result of normative assumptions (12, 22, 25, 28, 35). This was viewed as increasing their workload and involving longer appointment times (25, 35), and was associated with lower professional status (22) and overall, decreased satisfaction (12).

## Discussion

### Summary of findings

This review highlights barriers at personal, socio-cultural and system levels that inhibit women GPs’ capabilities, opportunities and motivations, leading to reduced career progression. While some positive changes have been documented across studies that span some thirty years, many challenges remain.

Most frequently these relate to historically gendered roles in the home and the associated challenges of childcare responsibilities and flexible working. Wider barriers due to medical cultures also appear slow to change; accounts of discriminatory and prejudiced behaviours are still alarmingly common (12, 25, 43).

Financial constraints were described, both in terms of women’s lower comparative earnings, financial pressures associated with maternity leave and women’s lower willingness to negotiate pay. Practices as employers should foster an environment where women feel comfortable discussing and negotiating pay with colleagues, and with standardised partner contracts that offer greater financial security during periods of maternity leave. While the New to Partnership Payment Scheme was introduced in 2020 to provide financial incentive and training to support greater uptake of partnership roles in general practice (44), knowledge of this scheme remains low and the financial incentive of £20,000 to new partners has been described as insufficient amidst wider uncertainty within the profession (25).

No evaluations of interventions to support women GPs’ career progression were identified in this review and there was a general lack of recent evidence which needs to be addressed. This is particularly important given the ongoing issues of GP wellbeing and retention, with evidence highlighting a differential impact on women GPs’ wellbeing across international studies (45).

### Strengths and limitations

Though our research focused on the experiences of UK doctors in general practice, findings are likely to translate to wider settings, both in family practice internationally and wider medical cultures. Our findings replicate those from specialities with historically lower proportions of women doctors (10, 46), highlighting wider societal and medical cultural challenges for women doctors.

Included studies were generally of good quality, though cross-sectional studies tended not to adjust for confounders and six studies sampled women only, which removes the ability to really investigate gender differences.

To our knowledge, this is the first systematic review of UK literature on this topic and the systematic approaches utilised throughout strengthens our findings. While all contributing authors were women, we engaged academic and medical doctors, which aided our interpretation of findings. Across the included studies gender was approached as binary with limited acknowledgement of the voices of those identifying as non-binary - a potential limitation of literature in this field at present.

### Implications for practice, policy, and future research

This review reveals a general consensus that general practice must adapt to become more flexible, supportive and balanced in terms of workload and leadership roles, in order to foster an environment where women can progress in their careers. While no intervention studies exist at present, through mapping the barriers women face to the COM-B model, this evidence synthesis may support the development of future policy initiatives to encourage greater participation in senior roles. Capability and Opportunity are described as acting as ‘logic gates’ by West and Michie, authors of the COM-B model (20), whereby *“both of the ‘gates’ (capability and opportunity) need to be open for motivation to generate the behaviour.”* Viewed quantitatively, this theory suggests the more women experiencing greater capability and opportunity over time will fuel future women’s motivation and potential to progress in their careers (20).

Areas for policy focus and evaluation may include improved flexibility in contracts, standardisation of partnership contractual conditions including maternity leave arrangements and access to childcare. Meanwhile mentorship schemes may reduce socio-cultural barriers through role modelling and supportive environments. Evaluation of all such schemes is required.

## Conclusion

Despite general practice being one of the more female-dominated medical specialties, barriers at personal, socio-cultural and system levels continue to inhibit women GPs’ careers.

## Declarations

### Ethical approval

This study did not require ethical or research governance approval.

### Author contributions

LJ designed and led the conduct of the review, with methodological expertise from SG and KB. SG designed the search strategy, undertook database searching and screened articles for inclusion, along with LJ, EW, KB and NG, who also undertook data extraction. LJ and EW synthesized findings and completed quality appraisal, with arbitration by VD. VD synthesized the quantitative data around study population characteristics. LJ and EW wrote the first draft of the article, to which all authors contributed. All authors have read and agreed the final version.

### Guarantor

LJ is study guarantor.

## Data Availability

All data produced in the present study are available upon reasonable request to the authors

