## Supplementary File for "Barriers and facilitators to women general practitioners’ career progression: a systematic review"

### Supplementary file A

#### Search strategy

**Database:**
Ovid MEDLINE(R) and Epub Ahead of Print, In-Process, In-Data-Review & Other Non-Indexed Citations and Daily <1946 to January 04, 2022>

| # | Query | Results from 5 Jan 2022 |
|---|---|---|
| 1 | exp United Kingdom/ | 381,776 |
| 2 | (national health service* or nhs*).ti,ab,in. | 234,887 |
| 3 | (english not ((published or publication* or translat* or written or language* or speak* or literature or citation*) adj5 english)).ti,ab. | 42,943 |
| 4 | (gb or "g.b." or britain* or (british* not "british columbia") or uk or "u.k." or united kingdom* or (england* not "new england") or northern ireland* or northern irish* or scotland* or scottish* or ((wales or "south wales") not "new south wales") or welsh*).ti,ab,jw,in. | 2,260,749 |
| 5 | (bath or "bath's" or ((birmingham not alabama*) or ("birmingham's" not alabama*) or bradford or "bradford's" or brighton or "brighton's" or bristol or "bristol's" or carlisle* or "carlisle's" or (cambridge not (massachusetts* or boston* or harvard*)) or ("cambridge's" not (massachusetts* or boston* or harvard*)) or (canterbury not zealand*) or ("canterbury's" not zealand*) or chelmsford or "chelmsford's" or chester or "chester's" or chichester or "chichester's" or coventry or "coventry's" or derby or "derby's" or (durham not (carolina* or nc)) or ("durham's" not (carolina* or nc)) or ely or "ely's" or exeter or "exeter's" or gloucester or "gloucester's" or hereford or "hereford's" or hull or "hull's" or lancaster or "lancaster's" or leeds* or leicester or "leicester's" or (lincoln not nebraska*) or ("lincoln's" not nebraska*) or (liverpool not (new south wales* or nsw)) or ("liverpool's" not (new south wales* or nsw)) or ((london not (ontario* or ont or toronto*)) or ("london's" not (ontario* or ont or toronto*)) or manchester or "manchester's" or (newcastle not (new south wales* or nsw)) or ("newcastle's" not (new south wales* or nsw)) or norwich or "norwich's" or nottingham or "nottingham's" or oxford or "oxford's" or peterborough or "peterborough's" or plymouth or "plymouth's" or portsmouth or "portsmouth's" or preston or "preston's" or ripon or "ripon's" or salford or "salford's" or salisbury or "salisbury's" or sheffield or "sheffield's" or southampton or "southampton's" or st albans or stoke or "stoke's" or sunderland or "sunderland's" or truro or "truro's" or wakefield or "wakefield's" or wells or westminster or "westminster's" or winchester or "winchester's" or wolverhampton or "wolverhampton's" or (worcester not (massachusetts* or boston* or harvard*)) or ("worcester's" not (massachusetts* or boston* or harvard*)) or (york not ("new york*" or ny or ontario* or ont or toronto*)) or ("york's" not ("new york*" or ny or ontario* or ont or toronto*))))).ti,ab,in. | 1,578,450 |
| 6 | (bangor or "bangor's" or cardiff or "cardiff's" or newport or "newport's" or st asaph or "st asaph's" or st davids or swansea or "swansea's").ti,ab,in. | 62,901 |
| 7 | (aberdeen or "aberdeen's" or dundee or "dundee's" or edinburgh or "edinburgh's" or glasgow or "glasgow's" or inverness or (perth not australia*) or ("perth's" not australia*) or stirling or "stirling's").ti,ab,in. | 233,035 |
| 8 | (armagh or "armagh's" or belfast or "belfast's" or lisburn or "lisburn's" or londonderry or "londonderry's" or derry or "derry's" or newry or "newry's").ti,ab,in. | 29,996 |
| 9 | or/1-8 | 2,839,068 |
| 10 | (exp africa/ or exp americas/ or exp antarctic regions/ or exp arctic regions/ or exp asia/ or exp australia/ or exp oceania/) not (exp United Kingdom/ or europe/) | 3,138,574 |
| 11 | 9 not 10 | 2,693,544 |
| 12 | physicians, women/ | 6,783 |
| 13 | ((female$ or women) adj2 (gp or gps)).mp. | 521 |
| 14 | ((female$ or women or gender) adj3 (family or general or primary) adj2 (doctor? or medicine or medical practitioner? or medical practice? or practice? or practitioner? or physician? or care or healthcare or health care)).mp. | 2,725 |
| 15 | career choice/ | 24,604 |
| 16 | Career Mobility/ | 11,888 |
| 17 | (career? or partner$ or principal or position or senior or salaried or portfolio).mp. | 937,505 |
| 18 | job satisfaction/ | 26,959 |
| 19 | health workforce/ | 13,973 |
| 20 | General Practitioners/og, sn, sd, td [Organization & Administration, Statistics & Numerical Data, Supply & Distribution, Trends] | 1,749 |
| 21 | (career? or partner$ or principal or position or senior or salaried or portfolio or self-employed).mp. | 938,875 |
| 22 | staff development/ | 9,776 |
| 23 | leadership/ | 44,751 |
| 24 | professional family relations/ | 15,371 |
| 25 | work-life balance/ | 944 |
| 26 | workforce.mp. | 101,768 |
| 27 | or/12-14 | 9,703 |
| 28 | or/15-26 | 1,102,331 |

| | | |
|---|---|---|
| 29 | 11 and 27 and 28 | 374 |

**Database:**
Embase <1974 to 2022 January 04>

| # | Query | Results from 5 Jan 2022 |
|---|---|---|
| 1 | exp United Kingdom/ | 438,442 |
| 2 | (national health service* or nhs*).ti,ab,in,ad. | 407,026 |
| 3 | (english not ((published or publication* or translat* or written or language* or speak* or literature or citation*) adj5 english)).ti,ab. | 50,730 |
| 4 | (gb or "g.b." or britain* or (british* not "british columbia") or uk or "u.k." or united kingdom* or (england* not "new england") or northern ireland* or northern irish* or scotland* or scottish* or ((wales or "south wales") not "new south wales") or welsh*).ti,ab,jx,in,ad. | 3,421,828 |
| 5 | (bath or "bath's" or ((birmingham not alabama*) or ("birmingham's" not alabama*) or bradford or "bradford's" or brighton or "brighton's" or bristol or "bristol's" or carlisle* or "carlisle's" or (cambridge not (massachusetts* or boston* or harvard*)) or ("cambridge's" not (massachusetts* or boston* or harvard*)) or (canterbury not zealand*) or ("canterbury's" not zealand*) or chelmsford or "chelmsford's" or chester or "chester's" or chichester or "chichester's" or coventry or "coventry's" or derby or "derby's" or (durham not (carolina* or nc)) or ("durham's" not (carolina* or nc)) or ely or "ely's" or exeter or "exeter's" or gloucester or "gloucester's" or hereford or "hereford's" or hull or "hull's" or lancaster or "lancaster's" or leeds* or leicester or "leicester's" or (lincoln not nebraska*) or ("lincoln's" not nebraska*) or (liverpool not (new south wales* or nsw)) or ("liverpool's" not (new south wales* or nsw)) or ((london not (ontario* or ont or toronto*)) or ("london's" not (ontario* or ont or toronto*)) or manchester or "manchester's" or (newcastle not (new south wales* or nsw)) or ("newcastle's" not (new south wales* or nsw)) or norwich or "norwich's" or nottingham or "nottingham's" or oxford or "oxford's" or peterborough or "peterborough's" or plymouth or "plymouth's" or portsmouth or "portsmouth's" or preston or "preston's" or ripon or "ripon's" or salford or "salford's" or salisbury or "salisbury's" or sheffield or "sheffield's" or southampton or "southampton's" or st albans or stoke or "stoke's" or sunderland or "sunderland's" or truro or "truro's" or wakefield or "wakefield's" or wells or westminster or "westminster's" or winchester or "winchester's" or wolverhampton or "wolverhampton's" or (worcester not (massachusetts* or boston* or harvard*)) or ("worcester's" not (massachusetts* or boston* or harvard*)) or (york not ("new york*" or ny or ontario* or ont or toronto*)) or ("york's" not ("new york*" or ny or ontario* or ont or toronto*))))).ti,ab,in,ad. | 2,659,098 |
| 6 | (bangor or "bangor's" or cardiff or "cardiff's" or newport or "newport's" or st asaph or "st asaph's" or st davids or swansea or "swansea's").ti,ab,in,ad. | 109,006 |
| 7 | (aberdeen or "aberdeen's" or dundee or "dundee's" or edinburgh or "edinburgh's" or glasgow or "glasgow's" or inverness or (perth not australia*) or ("perth's" not australia*) or stirling or "stirling's").ti,ab,in,ad. | 365,580 |
| 8 | (armagh or "armagh's" or belfast or "belfast's" or lisburn or "lisburn's" or londonderry or "londonderry's" or derry or "derry's" or newry or "newry's").ti,ab,in,ad. | 50,140 |
| 9 | or/1-8 | 4,178,441 |
| 10 | (exp "arctic and antarctic"/ or exp oceanic regions/ or exp western hemisphere/ or exp africa/ or exp asia/) not (exp united kingdom/ or europe/) | 3,203,266 |
| 11 | 9 not 10 | 3,952,704 |
| 12 | female physician/ | 5,246 |
| 13 | ((female$ or women) adj2 (gp or gps)).mp. | 695 |
| 14 | ((female$ or women or gender) adj3 (family or general or primary) adj2 (doctor? or medicine or medical practitioner? or medical practice? or practice? or practitioner? or physician? or care or healthcare or health care)).mp. | 3,338 |
| 15 | career planning/ | 3,248 |
| 16 | career/ | 27,277 |
| 17 | career mobility/ | 10,760 |
| 18 | (career? or partner$ or principal or position or senior or salaried or portfolio).mp. | 1,171,329 |
| 19 | job satisfaction/ | 32,618 |
| 20 | health workforce/ | 2,213 |
| 21 | (career? or partner$ or principal or position or senior or salaried or portfolio or self-employed).mp. | 1,172,989 |
| 22 | leadership/ | 77,199 |
| 23 | work-life balance/ | 2,116 |
| 24 | staff training/ | 14,708 |
| 25 | workforce.mp. | 38,813 |
| 26 | personnel management/ | 58,752 |
| 27 | physician shortage/ | 72 |
| 28 | skill mix/ | 428 |
| 29 | *general practitioners/ | 24,854 |
| 30 | or/12-14 | 8,985 |
| 31 | or/15-29 | 1,370,174 |
| 32 | 30 and 31 | 2,611 |

| | | |
|---|---|---|
| 33 | 11 and 32 | 372 |

**Database:**
HMIC Health Management Information Consortium <1979 to November 2021>

| # | Query | Results from 16 Dec 2021 |
|---|---|---|
| 1 | women/ and exp general practitioners/ | 141 |
| 2 | ((female$ or women) adj2 (gp or gps)).mp. | 120 |
| 3 | ((female$ or women or gender) adj3 (family or general or primary) adj2 (doctor? or medicine or medical practitioner? or medical practice? or practice? or practitioner? or physician? or care or healthcare or health care)).mp. | 203 |
| 4 | or/1-3 | 396 |
| 5 | exp careers/ | 417 |
| 6 | Career development/ | 595 |
| 7 | Career guidance/ | 108 |
| 8 | Career opportunities/ | 414 |
| 9 | Career patterns/ | 262 |
| 10 | career plans/ | 103 |
| 11 | (career? or partner$ or principal or position or senior or salaried or portfolio).mp. | 24,694 |
| 12 | job satisfaction/ | 1,131 |
| 13 | Staff morale/ | 291 |
| 14 | workforce/ | 4,971 |
| 15 | (career? or partner$ or principal or position or senior or salaried or portfolio or self-employed).mp. | 24,884 |
| 16 | Human resources development/ | 376 |
| 17 | exp Leadership/ | 3,283 |
| 18 | exp Leave/ or exp Staff working life/ or exp Flexible working hours/ or exp Working hours/ or exp Flexible working/ | 2,544 |
| 19 | workforce.mp. | 9,151 |
| 20 | exp Skill mix/ | 615 |
| 21 | exp Staff shortage/ | 766 |
| 22 | exp general practitioners/ | 10,334 |
| 23 | exp leave/ | 880 |
| 24 | Staff working life/ | 85 |
| 25 | Flexible working hours/ | 55 |
| 26 | exp Working hours/ | 1,400 |
| 27 | Flexible working/ | 291 |
| 28 | exp working conditions/ | 3,150 |
| 29 | staff retention/ | 1,586 |
| 30 | exp Family role/ | 38 |
| 31 | or/5-30 | 50,821 |
| 32 | 4 and 31 | 273 |

#### Google Scholar

To limit the number of records to sift we decided on an approach of either importing the first 100 records into Endnote or all results up to point where 10 irrelevant records are identified sequentially (whichever comes first).
Potentially relevant records were identified throughout the first 100 records. Therefore, the first 100 of about 4,680,000 search results were entered into the Endnote Library after using the following search strategy
(women OR female) AND (partners OR partnerships OR career OR principals OR work) AND (GP OR GPs OR "general practitioners" OR "primary care")
